# Impact of adverse event reporting system on burnout and job satisfaction of nurses in China: a nationwide cross-sectional study using a multilevel modeling approach

**DOI:** 10.1101/2022.09.13.22279881

**Authors:** Yanhua Chen, Yanrong He, Peicheng Wang, Feng Jiang, Yanrong Du, Ming Yen Cheung, Huanzhong Liu, Yuanli Liu, Tingfang Liu, Yi-lang Tang, Jiming Zhu

**Author notes:** **Corresponding author:** Jiming Zhu, Vanke School of Public Health, Tsinghua University, Beijing 100084, China. *E-mail address:.

## Abstract

**IMPORTANCE:** Many organizational and personal factors may contribute to burnout and poor job satisfaction experienced by nurses. Adverse Event Reporting System (AERS) is a key component of organizational-level quality improvement system which impacts workplace wellness for nurses. However, little is known about the underlying influence and mechanism that AERS have on nurse’ well-being.

**OBJECTIVE:** To explore the relationship between the implementation of AERS, burnout, and job satisfaction among psychiatric nurses in China. To uncover the mechanism through which AERS influences burnout and job satisfaction, while examining the mediating effect of workplace violence from patients.

**DESIGN, SETTING, AND PARTICIPANTS:** This cross-sectional study used the multilevel linear regression analyses with a national sample of 9,744 psychiatric nurses from 41 psychiatric hospitals across 29 provinces in China. Data collection was performed in March 2019, and the analyses were conducted from May to July 2022.

**MAIN OUTCOMES AND MEASURES:** The main outcomes were self-reported burnout and job satisfaction. Burnout was measured by the Maslach Burnout Inventory and job satisfaction was measured using Minnesota Satisfaction Questionnaire.

**RESULTS:** Among 9,744 nurses (mean [SD] age, 34.7 [8.8] years) interviewed, 8064 (82.8%) were female. AERS was positively associated with job satisfaction (β=3.70; p<0.05), but negatively with burnout (β=-3.42; p<0.01) and workplace violence (β=-0.55; p<0.1). Workplace violence was positively associated with burnout (β=2.27; p< 0.01), while negatively associated with job satisfaction (β=-0.81; p<0.01). Mediation analysis indicated that the association between AERS, burnout, and job satisfaction was mediated by workplace violence.

**CONCLUSIONS AND RELEVANCE:** This study highlights that the implementation of AERS is associated with less workplace violence in hospitals, and it may also contribute to lower levels of burnout and higher levels of job satisfaction among psychiatric nurses.

**Key points:** *Question:* How does hospital’s Adverse Event Reporting System (AERS) impact nurses’ well-being? What are the underlying mechanisms?

*Findings:* In this cross-sectional study of 9744 psychiatric nurses, nurses in hospitals with AERS reported significantly lower burnout and job dissatisfaction than those in hospitals without AERS. AERS seems to impact well-being by reducing workplace violence.

*Meaning:* Although Chinese authorities recently stipulated AERS, not all healthcare organizations have fully implemented it. Reporting and consequent quality improvement actions seem to alleviate workplace violence, especially in patient-provider relationship. This will inform hospital management to further leverage AERS for workplace well-being and satisfaction.

## INTRODUCTION

Burnout and poor well-being have been recognized as common occupational hazards among physicians, nurses, and other healthcare professionals^1,2,3^. Nurse burnout and poor well-being are found to be associated with job turnover^4,5^, medical errors^6^, and poor patient care^7,8^. Many factors including individual, institutional and social factors may contribute to burnout and poor well-being in healthcare professionals, therefore multi-level interventions are often needed. Individual-focused approaches such as those aimed at mental resilience have been taken to reduce distress and promote well-being among nurses^9^. Institutional or organizational factors are also important. Since burnout and low job satisfaction are occupational problems and may potentially influence health quality, healthcare facility management needs to ensure a positive and healthy workplace^10,11^.

Workplace violence from patients is an increasing threat to the safety and well-being of healthcare workers in China. In recent years, high rates of workplace violence against healthcare professionals have often been reported in different settings^12,13^. In cases of serious violence, there were murders and injuries, such as a doctor who had acid poured on his face and another whose throat was cut^14^. A national survey showed that 82% of psychiatric nurses reported encountering either physical or verbal assault during the past year^15^. Further studies demonstrated that as violence increased, healthcare providers often experienced higher levels of burnout and lower levels of well-being^16,17^. The violence initiated by patients and their relatives is at least partially caused by poor healthcare quality^18^. Accordingly, minimizing workplace violence could establish a safe and healthy workplace, which would be a potential solution to burnout and job dissatisfaction. A specific strategy for improving healthcare quality has the potential to promote well-being.

Adverse events negatively affect the quality of medical care^19^ and contribute to undesirable harm in patients due to medical errors or improper management^20^. Investigation of adverse events provides information on incidences that can be used to identify areas of risk and to inform amenable actions^21,22^. In 2011, a new policy of medical quality and security incidents reporting was launched by the China’s Ministry of Health^23^. The policy defined list of information for hospitals to report to health administration while investigating the cause to prevent it from occurring again. Adverse Event Reporting System (AERS) is a hospital information system that farthest facilitates the collecting of adverse event data and supports investigation of data for quality improvement^24^. In China, some hospitals have adopted AERS that consists of functional modules such as Event Reporting, Event Inquiry, Statistical Inquiry, Statistical Analysis, Authority Management, and System Configuration^25^. For hospital management, the reporting data enables them to analyze the root causes of adverse events and prevent recurrence^26^. AERS also brings indirect benefits. Health professionals become “secondary victims” when patients experience adverse events^27,28^. From this perspective, AERS is a strategy to improve healthcare quality at the organizational level to address a safe and healthy workplace for nurses. It can promote providers’ well-being by preventing medical errors and workplace violence^29,30,31^.

Existing literature on AERS has paid much attention to factors that impede healthcare providers from reporting adverse events^32,33,34^, while ignoring that AERS may promote a healthy workplace and boost nurses’ occupational well-being. To fill these gaps, our study aims to investigate the impact of AERS on burnout and job satisfaction of nurses based on data from a national survey of psychiatric hospitals in China. We further explore the possible channels through which AERS reduces burnout and drives higher job satisfaction for nurses in China by analyzing the mediating role of workplace violence.

## METHODS

### Study design and Participants

A cross-sectional study was conducted in 41 psychiatric hospitals across China. All nurses working in the sampled hospitals were invited. A self-administered questionnaire with two-level data was distributed among healthcare workers and hospital managers. Personal information, burnout, job satisfaction, and physical health status were asked for psychiatric nurses.

Information about the AERS was collected from the hospital managers. This cross-sectional study followed the Strengthening the Reporting of Observational Studies in Epidemiology (STROBE) reporting guideline.

The study was part of the National Hospital Performance Evaluation Survey (NHPES) conducted in March 2019. The survey aimed to cover all 31 provinces in Mainland China except two provinces, Gansu and Tibet, as these two had no psychiatric tertiary hospitals. Accordingly, 41 psychiatric tertiary hospitals from 29 provinces in China were included in the study. The sample was representative by geographical area (eFigure 1 in the Supplement): 14 hospitals in Eastern China, 9 hospitals in Central China, 12 hospitals in Western China, and 6 hospitals in Northeastern China. The questionnaire was sent to healthcare workers by Wechat anonymously, which is a widely and frequently used app for instant messaging and social interaction.

The study was approved by the Ethics Committee of Chaohu Hospital of Anhui Medical University (No. 201903-kyxm-02) before initiation. Informed consent was obtained from participants before they started the questionnaire.

### Measures

AERS is a binary hospital-level variable (1 for hospitals that have an adverse event reporting system in place, and 0 otherwise). AERS is one of the questions concerning healthcare quality management approaches answered by hospital managers. Respondents working at hospitals with AERS will be further asked to provide the name of the system.

Workplace violence was measured by two questions concerning the verbal and physical violence participants had experienced in the workplace from patients. The first question is “How many times, in the past 12 months, did you find yourself in a situation of verbal aggression (e.g., expressions of abuse, slandering, contempt, insulting, or humiliating without physical contact) by patients?”. The second question is “How many times, in the past 12 months, did you find yourself in a situation of physical aggression by patients (e.g., pushing, hitting, inflicting, and physical harm on persons or violence with weapons)?” Answers are scored as: 1 = never/almost never, 2 = <12 times/year, 3 = once a month, 4 = 2-3 times/month, 5 = once a week, 6 = 2-5 times/week, 7 = almost every day. The prevalence of experienced workplace violence in this study was the sum of verbal and physical violence. The intra-class correlation (ICC) of workplace violence is 0.077.

Burnout was assessed using the Maslach Burnout Inventory (MBI) scale, a widely used scale to assess burnout^35^ which has been used in Chinese samples^36,37^. The subscales include emotional exhaustion (EE, 9 items), depersonalization (DP, 5 items), and personal accomplishment (PA, 8 items). All items are scored on a seven-point Likert scale, ranging from 0 (never) to 6 (every day). In this study, a high degree of burnout is composed of high levels of EE and DP (ranging from 0 to 84)^38,39^, with Cronbach’s alpha being 0.8864. Aggregation of the data at the hospital level was justified (ICC=0.051).

Job satisfaction was measured using the 20-item Minnesota Satisfaction Questionnaire (MSQ). The Chinese version has been widely used and has demonstrated good reliability and validity^40^. All items were answered on a 5-point Likert scale, ranging from 1 for “very unsatisfied” to 5 for “very satisfied”. The total MSQ score (ranging from 20 to 100) was calculated to indicate the extent of job satisfaction among healthcare workers. A higher score indicates a higher level of satisfaction with their job. Cronbach’s alpha was 0.9557. Aggregation of the data at the hospital level was justified (ICC=0.129).

To adjust for regression models, hospital characteristics and nurses’ characteristics were taken into account as covariates based on the theoretical causal pathways between AERS and well-being. Hospital-level potential confounders selected from NHPES included: the number of available beds used to capture the hospital size, the outpatient number applied to account for service provision, and the amount of training for patient safety employed to express the emphasis on patient safety in the hospital. Each of them was transformed by quartile, i.e., 25_th_, 50_th_, and 75_th_ percentile of the number. Individual-level factors included age, gender (male or female), an education level (associate degree or lower, bachelor’s degree, or master’s degree or higher), monthly after-tax income (less than 5000, 5001-8000, 8001-12000, or more than 12000, in Chinese Yuan), and night shift during the past month.

### Statistical analysis

Descriptive statistics such as frequencies, percentages, means, and standard deviations, were given in Table 1. Multilevel linear regression analyses were carried out to examine if AERS impacts burnout and job satisfaction, and to identify the impact of workplace violence. Associations between AERS, workplace violence, burnout as well as job satisfaction were examined after adjusting for hospital and sociodemographic confounders. The median standardized *β* and standard error (SE) were reported for all model specifications. To evaluate the mediating role of workplace violence, we used the procedure proposed by Baron and Kenny^41^. The mediating effects exist when the independent variable (AERS) has a significant effect on the mediating variable (workplace violence). The mediating variable also has significant effects on the dependent variables (burnout and job satisfaction). In addition, Sobel tests were used to assess the significance of the mediating effects^42^. We used 0.1, 0.05, and 0.01 as the cutoff points for the significant levels. All analyses were done with STATA, version 17.

**Table 1.** Demographic characteristics of the participants

| Characteristic | Total participants<br>(n = 9744) | Hospitals with AERS<br>(n = 7045) | Hospitals without AERS<br>(n = 2699) |
| --- | --- | --- | --- |
| Age, mean (SD), years | 34.7 (8.8) | 34.91 (8.81) | 34.22 (8.68) |
| Gender |  |  |  |
| Male | 1680 (17.2) | 1236 (17.54) | 444 (16.45) |
| Female | 8064 (82.8) | 5809 (82.46) | 2255 (83.55) |
| Education level |  |  |  |
| Associate degree or less | 3361 (34.5) | 2229 (31.64) | 1132 (41.94) |
| Bachelor's degree | 6323 (64.9) | 4766 (67.65) | 1557 (57.69) |
| Master's degree or above | 60 (0.6) | 50 (0.71) | 10 (0.37) |
| Working years |  |  |  |
| <5 | 1911 (19.6) | 1316 (18.68) | 595 (22.05) |
| 5-9 | 2564 (26.3) | 1875 (26.61) | 689 (25.53) |
| 10-20 | 2756 (28.3) | 2001 (28.40) | 755 (27.97) |
| >20 | 2513 (25.8) | 1853 (26.30) | 660 (24.45) |
| Monthly income after tax<sup>a</sup> |  |  |  |
| Low: ≤5000 RMB* | 4237 (43.5) | 2890 (41.02) | 1347 (49.91) |
| Medium: 5001-8000 RMB | 3463 (35.5) | 2657 (37.71) | 806 (29.86) |
| High 8001-12000 RMB | 1732 (17.8) | 1272 (18.06) | 460 (17.04) |
| Very high >12000 RMB | 312 (3.2) | 226 (3.21) | 86 (3.19) |
| Night shift during the past month,<br>mean (SD), days | 4.46 (4.2) |  |  |
| ≤ 4 | 5148 (52.8) | 3660 (51.95) | 1488 (55.13) |
| >5 | 4596 (47.2) | 3385 (48.05) | 1211 (44.87) |

<sup>a</sup> 6.47 RMBs = 1 USD at time of the survey.

## RESULTS

### Descriptive analysis of hospitals and participants

Basic demographic and hospital information were collected in the NHPES. Of the 41 hospitals, the number of available beds ranged from 169 to 2141. The total number of outpatient visits ranged from 60,755 to 1,780,102 in the last two years. The amount of staff training sessions on healthcare quality improvement and patient safety provided by hospitals ranged from 2 to 145 during the past two years. Of the 41 hospitals, 28 (68.3%) had AERS, whereas 13 (31.7%) had no AERS. More detailed information on difference between AERS and non-AERS hospitals were reported in eTable 1 in the Supplement.

A total of 13,867 psychiatric nurses were invited to participate, and 9,744 of them completed the questionnaire (response rate was 70.3%). Table 1 summarized the characteristics of the participants. In this study, the average age of the participants was 34.7 years old, and most of them were female (82.8%). More than half of the participants had a bachelor’s degree (64.9%). 19.6% of the participants had worked less than 5 years, and more than half (64.8%) of them worked less than 40 hours per week. In terms of the monthly income, 43.5% of nurses earned less than 5000 RMBs (approximately $773 USD). The average number of night shifts during the past month was 4.46.

### Prevalence of burnout and job satisfaction in different settings

As shown in figure 1, nurses in hospitals with AERS reported significantly lower burnout than those in hospitals without AERS (25.53 vs.28.13, p<0.01; Figure. 1A). Similarly, nurses in hospitals with AERS reported significantly higher job satisfaction than those in hospitals without AERS (68.93 vs 65.97, p<0.01; Figure. 1B). Workplace violence among nurses in hospitals with AERS was less prevalent than it in hospitals without AERS (5.54 vs. 6.10, p<0.01; Figure. 1C). Details for each hospital were summarized in eTable 2 in the Supplement.

**Figure 1.**
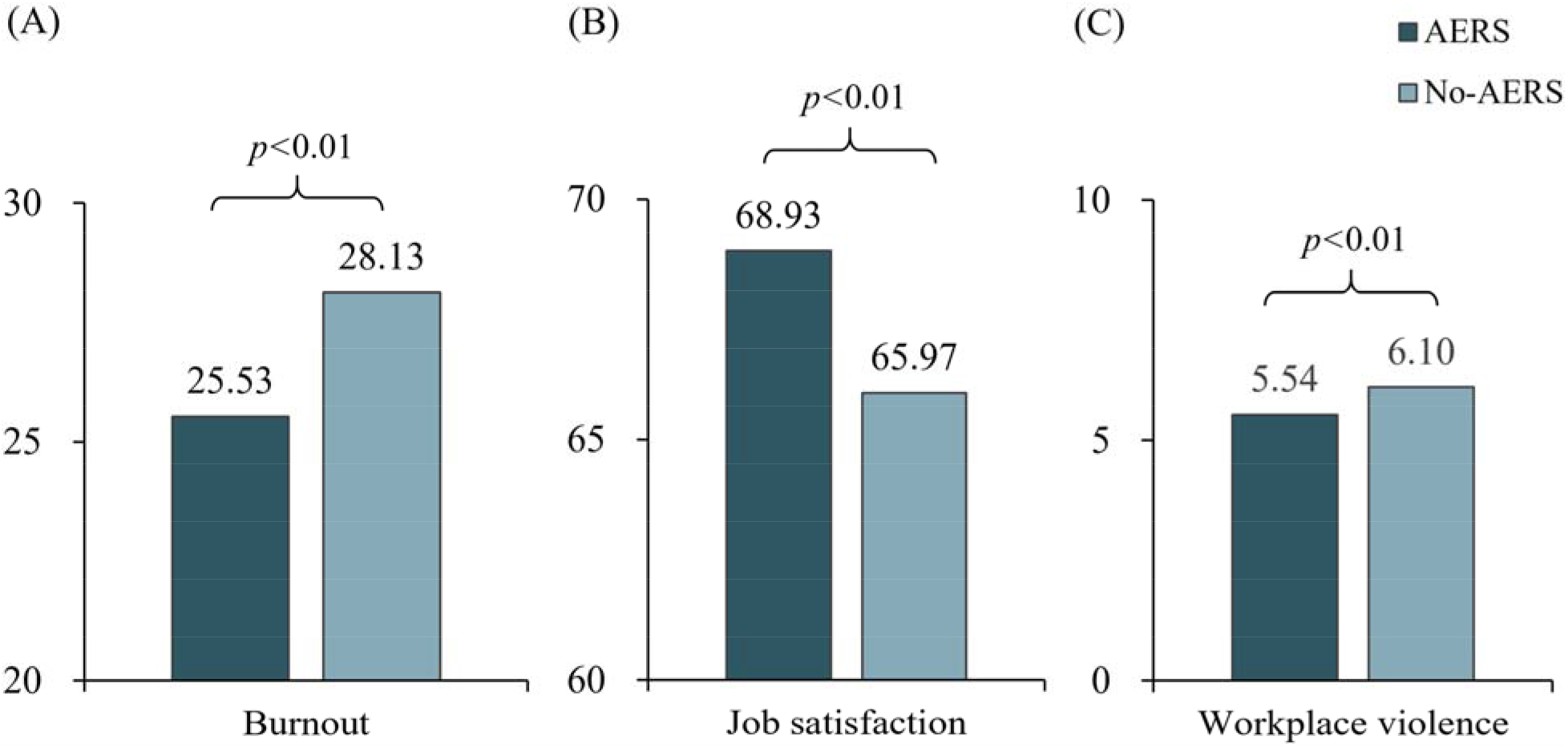
Difference analysis of burnout, job satisfaction and workplace violence in settings. The left three bars show the mean value of burnout, job satisfaction, and workplace violence in hospitals with AERS and the others without AERS.

### Factors associated with burnout and job satisfaction

Table 2 presents estimated coefficients of the relationships between AERS, burnout, and job satisfaction using multilevel regression analyses. Results of model 1 and model 2 indicate that nurses in the hospital with AERS have lower levels of burnout (β=-3.42; p<0.01) and a higher level of job satisfaction (β=3.70; p<0.05), after adjusting for other variables.

**Table 2.**
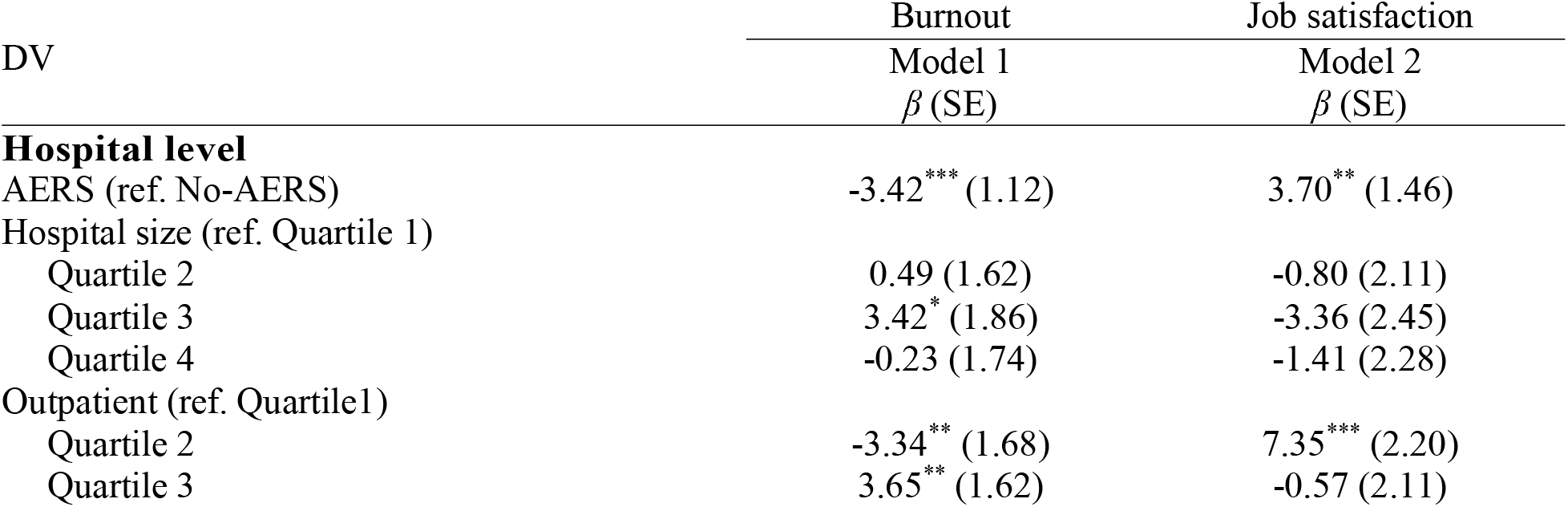
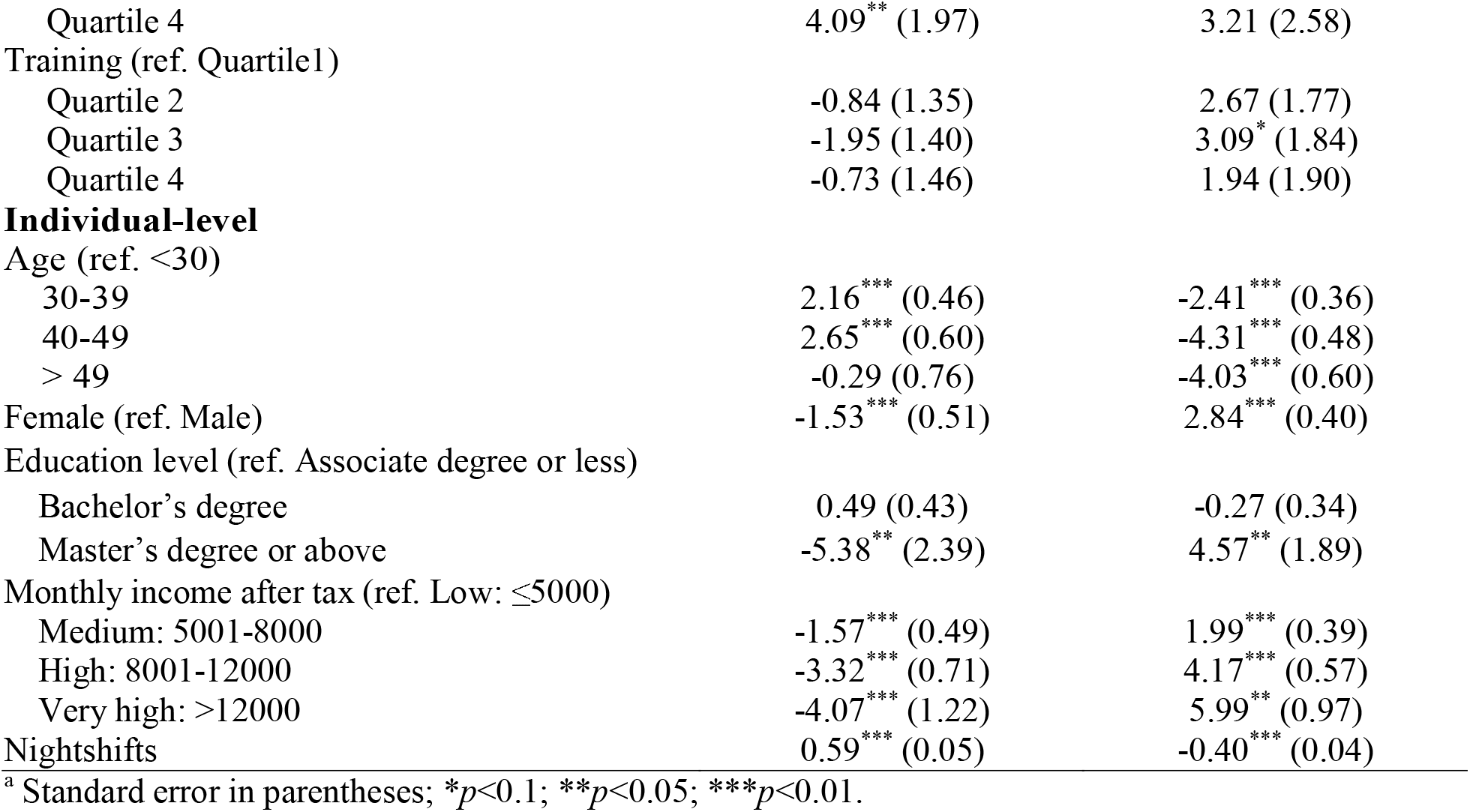
Multilevel analyses for AERS associated with burnout and job satisfaction ^a^

### The mediating role of workplace violence

Table 3 shows the mediation effects of workplace violence using multilevel analyses. Hospital and individual level sociodemographic confounders are included in the models. Model 3 found a significantly negative association between AERS and workplace violence (β = -0.55; p<0.1). When AERS and workplace violence are set as predictors and burnout as the dependent variable in Model 4, workplace violence had a significantly negative effect on burnout (β = 2.27, p<0.01). When compared to the total effect (= -3.42, p<0.01), the direct effect of AERS on burnout was reduced (= -2.14, p<0.05). Similarly, when job satisfaction was set as a dependent variable in Model 5, workplace violence was negatively associated with job satisfaction (β= -0.81, p<0.05), and the direct effect (β = 3.25, p<0.05) of AERS on job satisfaction was also reduced when compared with the total effect (β = 3.70, p<0.05). The decreased coefficient further confirmed the mediating role workplace violence played in the associations. The mediation analyses were represented as a path diagram in Figure 2. We examined the mediating role of verbal and physical violence respectively as sensitivity analyses shown in eTable 3-4 and eFigure 2-3 in the Supplement.

**Table 3.**
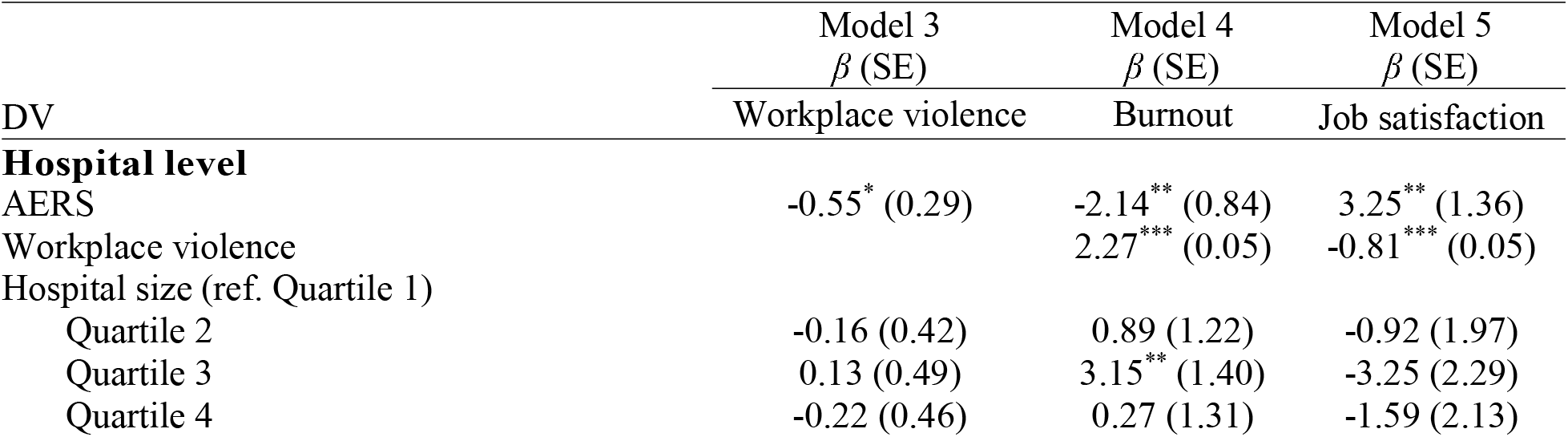
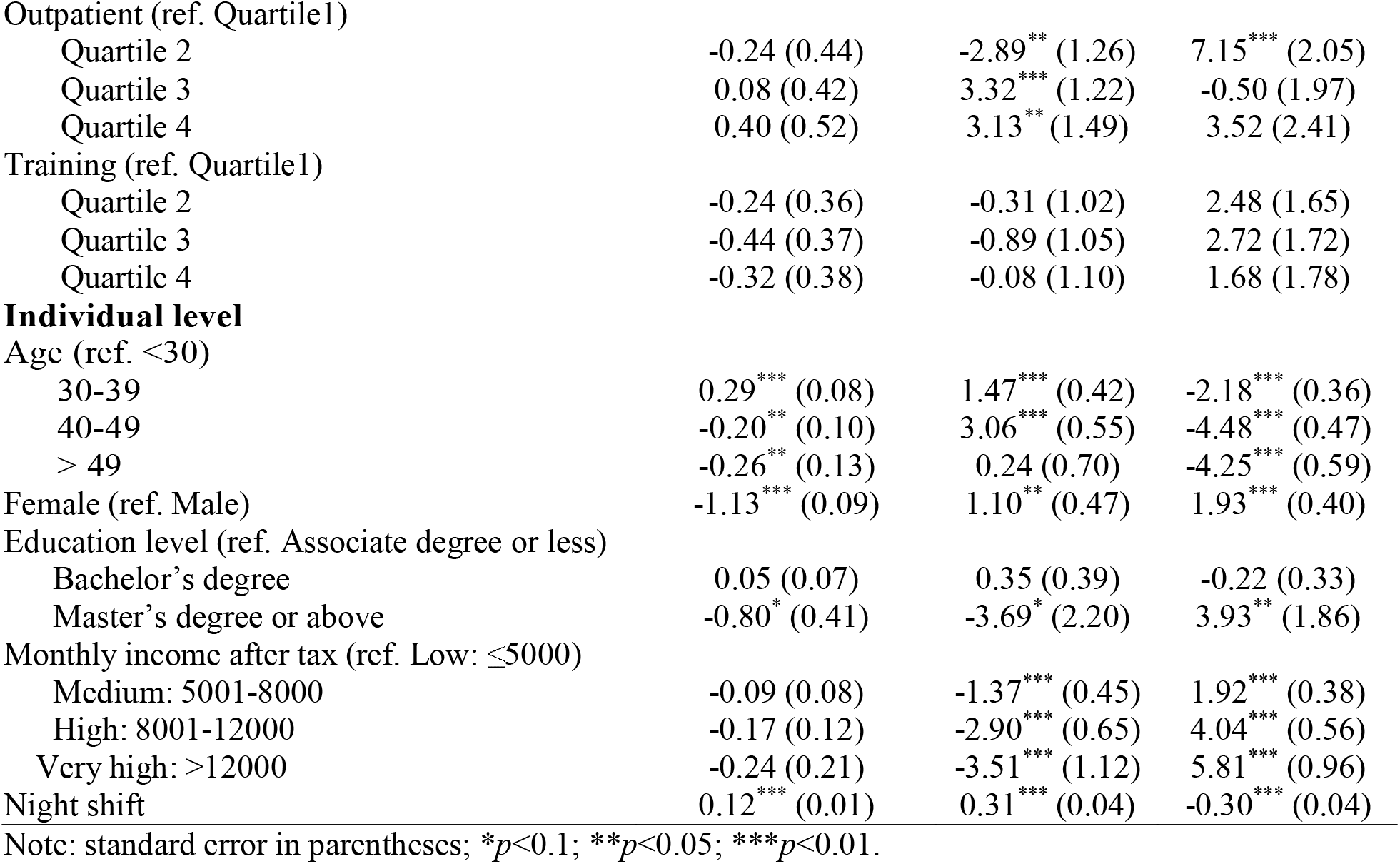
Relationship between workplace violence associated with burnout and job satisfaction

**Figure 2.**
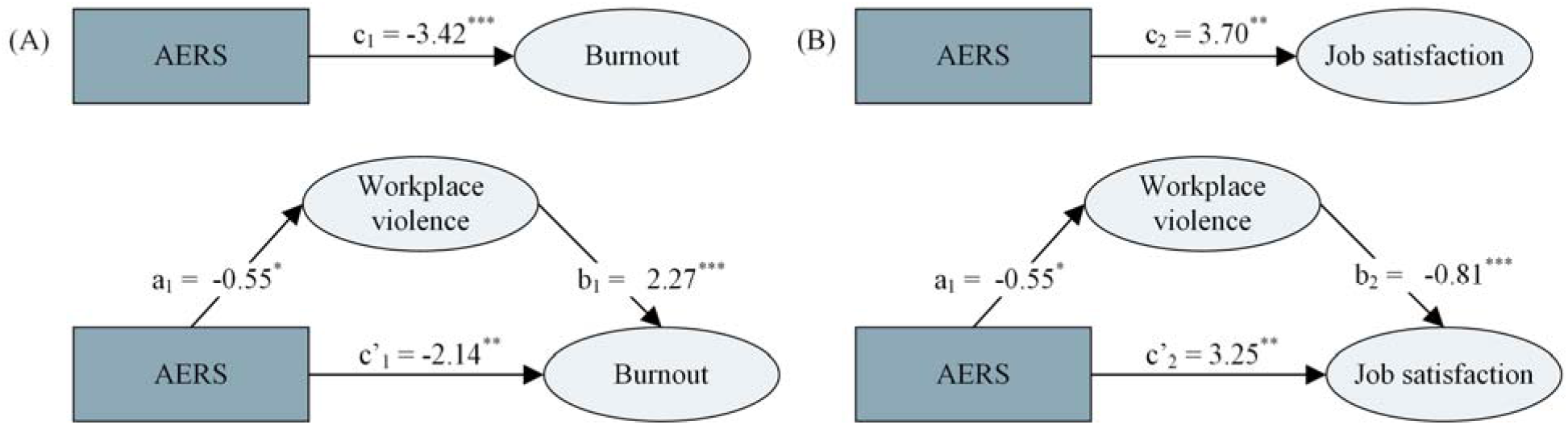
Mediation analysis. Path diagram shows that workplace violence was the mediator between AERS and burnout as well as job satisfaction. All values indicated were calculated as follows in model 1-5: Burnout = c_1_ × AERS + e_1_; Job satisfaction = c_2_ × AERS + e_2_; Workplace violence = a_1_ (a_2_) × AERS + e_3_; Burnout = c’_1_×AERS + b_1_×Workplace violence + e_4_; Job satisfaction = c’_2_ × AERS + b_2_×Workplace violence + e_5_. \**p*<0.1; \*\**p*<0.05; \*\*\**p*<0.01.

The Sobel tests provide additional support for the mediating effect of workplace violence on the relationship between AERS and burnout (Zsobel = -1.89; Std. Error = 0.66; P =0.06) as well as on the relationship between AERS and job satisfaction (Zsobel = 1.88; Std. Error = 0.24; P =0.06). In summary, workplace violence was negatively and significantly associated with AERS, suggesting that the implementation of AERS in the hospital is associated with a decreased prevalence of workplace violence against nurses at the individual level. The association of AERS with burnout and with job satisfaction was attenuated by workplace violence from patients and their family members.

## DISCUSSION

We analyzed data from a large national sample of nurses from 41 psychiatric hospitals in China using a multilevel modeling approach. We found a positive and significant association between AERS and nurses’ well-being. Compared to hospitals without AERS, nurses in hospitals with AERS reported significantly lower burnout and higher job satisfaction. These associations were consistent after adjusting for a wide range of confounding covariates at the hospital level (i.e. the hospital size, the outpatient number, and the amount of training for patient safety) and individual level. Moreover, we observed a mediating effect of workplace violence on the association between AERS and nurses’ well-being. Our findings provide evidence that AERS is an initiative to create positive work environments to reduce the risk of burnout and foster professional well-being^43^.

A considerable amount of work has been done to promote nurses’ awareness^44^ and knowledge^45^ of AERS, such as surveying their adoption willingness^46,47^ and perceived barriers^48^ toward voluntary reporting of adverse events. Nurses were found reluctance to report adverse events^49,50,51^. For instance, nurses may be afraid of negative consequences, and they may view the reporting system as a burden due to work pressure and insufficient time to report^52^. These perceptions of AERS vary by individual characteristics and personal experience. At organizational level, previous studies also revealed that the implementation of AERS would increase the rate^53^ and the number^54^ of reporting medical errors, and also decrease the error severity^55^. However, existing research has neglected the system-level improvement of implementing AERS, which influences all nurses in the workplace simultaneously and equivalently, regardless of their individual experiences. Existing research has neglected this influence. Our study addresses this knowledge gap using a multilevel modeling approach and provides new evidence for the system-level improvements in the work environment can reduce nurse burnout^56^. According to this study, though nurses were afraid of negative consequences and hesitant to report adverse events individually, AERS would reduce these concerns by addressing the issue systematically. The finding from our study informs hospital management to further leverage AERS for workplace improvement in other areas.

The current study found that AERS was significantly associated with decreased burnout and increased job satisfaction among psychiatric nurses. AERS was developed to primarily reduce adverse events^57,58^, and experiencing adverse events may contribute to burnout^59^, moral distress^60^, and job dissatisfaction^61^ in nurses, which could explain our findings about the associations between AERS and nurses’ well-being. Our finding can be explained by previous findings that the more helpful information system was perceived by healthcare providers, the more satisfied they were with their job^62^. In addition, these results are consistent with a previous finding that hospital nursing units with a greater number of adverse events had a higher rate of burnout among nurses^63^.

In the present study, the prevalence of workplace violence in hospitals with AERS was lower than in those without AERS, and there was a significantly negative association between AERS and workplace violence. The possible explanation is that AERS provides better channel for data collection and transparency that weren’t possible before as patient-provider tension is often exacerbated by mismatching information. Also, when the AERS is integrated with other hospital information systems such as electronic health record systems, drug management systems, and clinical decision support systems, this may enhance root cause finding and preventing patient care errors. AERS can also facilitates patient-reported adverse events data, which empowers patients and enhances patient safety^64^. Our finding indicates great potential in optimizing the information system and utilizing AERS in conjunction with quality improvement tools such as Continuous Quality Improvement, Lean Six Sigma Initiatives^65^. With the right tools, the management will be able to handle the incident promptly through risk assessment and follow-up action plans^66^.

Our results also showed that workplace violence played a mediating role in the association between AERS and nurse well-being. On the one hand, the results align well with previous findings that AERS could improve patient safety^67^, which is a key determinant of patient satisfaction^68^. Additionally, our findings are in accordance with numerous prior findings that workplace violence was associated with burnout and low job satisfaction among healthcare professionals^69,70,71^. Our findings provide fresh insights into how AERS improves nurse well-being. In other words, the implementation of AERS to prevent adverse events may reduce workplace violence between providers and patients, which would be beneficial to well-being. To amplify the positive effect of AERS, a non-punitive culture accompanied by supportive policies such as anonymous procedure, staff education, audit practice, and feedback procedures should be encouraged to eliminate the fear of being blamed and improve healthcare quality^72,73,74^.

Several limitations need to be acknowledged in this study. First, the survey only asked if AERS existed in the hospital or not. We did not collect data on the details about the specific functions of AERS. The functions performed by AERS may vary, which may provide more insights into individual module within AERS. Second, our findings and conclusions are derived from a survey of psychiatric hospitals. NHPES is the latest nationwide representative survey in psychiatric settings and reflects the current situation of psychiatrists. Our findings may not be generalizable to other healthcare facilities, such as general hospitals where workplace violence is less intensive. Future studies need to include different hospitals. Third, limited by the data resources, more mechanisms with other potential mediators (i.e., patient safety, care quality, medical errors, nurse-patient relationships, or job performance) need to be studied. Finally, our study is cross-sectional so causality cannot be inferred. We assume that the direction is from AERS to workplace violence and well-being. However, the reciprocal causation or third variable cannot be ruled out even by a multilevel research design. In addition, although we have controlled a range of covariates both at the hospital (organizational) and individual levels, there remains the possibility for unobserved biases. Due to the limited access to data, we didn’t consider other confounding factors (e.g., organizational culture, resource levels) that would influence both workplace violence and nurse well-being.

Despite these limitations, this study is, to our knowledge, the first to shed light on the association between AERS with burnout and job satisfaction. Our results demonstrate the importance of organizational strategies to reduce burnout and improve job satisfaction. Specifically, the application of AERS can improve individual well-being, regardless of their personal experience or perception of AERS. The system for adverse event reporting would also benefit the hospital overall. This study also verified the unintentionally favorable impact of an error reporting system through establishing a safe and healthy workplace.

## Supporting information

STROBE Statement

## Data Availability

All data produced in the present study are available upon reasonable request to the authors

## References

1 Tawfik, D. S., Profit, J., Morgenthaler, T. I., Satele, D. V., Sinsky, C. A., Dyrbye, L. N., … & Shanafelt, T. D. (2018, November). Physician burnout, well-being, and work unit safety grades in relationship to reported medical errors. In Mayo Clinic Proceedings (Vol. 93, No. 11, pp. 1571–1580). Elsevier.

2 Busis, N. A., Shanafelt, T. D., Keran, C. M., Levin, K. H., Schwarz, H. B., Molano, J. R., … & Cascino, T. L. (2017). Burnout, career satisfaction, and well-being among US neurologists in 2016. Neurology, 88(8), 797–808.

3 Bodenheimer, T., & Sinsky, C. (2014). From triple to quadruple aim: care of the patient requires care of the provider. The Annals of Family Medicine, 12(6), 573–576.

4 Shah, M. K., Gandrakota, N., Cimiotti, J. P., Ghose, N., Moore, M., & Ali, M. K. (2021). Prevalence of and factors associated with nurse burnout in the US. JAMA network open, 4(2), e2036469–e2036469.

5 Ran, L. et al., 2020. Job burnout and turnover intention among Chinese Primary Healthcare Staff: The mediating effect of satisfaction. BMJ Open, 10(10).

6 Hall, L.H. et al., 2016. Healthcare staff wellbeing, Burnout, and Patient Safety: A Systematic Review. PLOS ONE, 11(7).

7 Clark, R.R. & Lake, E., 2020. Burnout, job dissatisfaction and missed care among maternity nurses. Journal of Nursing Management, 28(8), pp.2001–2006.

8 Bodenheimer, T., & Sinsky, C. (2014). From triple to quadruple aim: care of the patient requires care of the provider. The Annals of Family Medicine, 12(6), 573–576.

9 West CP, Dyrbye LN, Sinsky C, et al. Resilience and Burnout Among Physicians and the General US Working Population. JAMA Netw Open. 2020;3(7):e209385.

10 Carayon, P., Cassel, C., & Dzau, V. J. (2019). Improving the system to support clinician well-being and provide better patient care. JAMA, 322(22), 2165–2166.

11 Montgomery, A., Panagopoulou, E., Esmail, A., Richards, T., & Maslach, C. (2019). Burnout in healthcare: the case for organisational change. Bmj, 366.

12 The Lancet (2020). Protecting Chinese doctors. Lancet 395:90. doi: 10.1016/s0140-6736(20)30003-9

13 Yang Q, Tai-Seale M, Liu S, et al. Measuring Public Reaction to Violence Against Doctors in China: Interrupted Time Series Analysis of Media Reports. J Med Internet Res 2021;23(2):e19651.

14 Hesketh T, Wu D, Mao L, et al. Violence against doctors in China. BMJ 2012;345:e5730.

15 Han, X., Jiang, F., Shen, L., Liu, Y., Liu, T., Liu, H., … & Zhu, J. (2022). Workplace violence, workforce stability, and well-being in China’s Psychiatric Hospitals. American journal of preventive medicine, 62(4), e265-e273.

16 Wu, Y., Jiang, F., Ma, J., Tang, Y.-L., Wang, M., and Liu, Y. (2021). Experience of medical disputes, medical disturbances, verbal and physical violence, and burnout among physicians in China. Frontiers in psychology, 3132.

17 Chen, Y., Wang, P., Zhao, L., He, Y., Chen, N., Liu, H., … & Zhu, J. (2022). Workplace violence and turnover intention among psychiatrists in a national sample in China: the mediating effects of mental health. Frontiers in psychiatry, 13.

18 Yu H, Hu Z, Zhang X, et al. How to overcome violence against Healthcare professionals, reduce medical disputes and ensure patient safety. Pak J Med Sci 2015;31(1):4–8.

19 Heavner, J. J., & Siner, J. M. (2015). Adverse event reporting and quality improvement in the intensive care unit. Clinics in chest medicine, 36(3), 461–467.

20 Voluntary Electronic Reporting of Medical Errors and Adverse EventsAn Analysis of 92,547 Reports from 26 Acute Care Hospitals. JOURNAL OF GENERAL INTERNAL MEDICINE

21 De Vries, E. N., Ramrattan, M. A., Smorenburg, S. M., Gouma, D. J., & Boermeester, M. A. (2008). The incidence and nature of in-hospital adverse events: a systematic review. BMJ Quality & Safety, 17(3), 216–223.

22 Rafter, N., Hickey, A., Condell, S., Conroy, R., O’connor, P., Vaughan, D., & Williams, D. (2015). Adverse events in healthcare: learning from mistakes. QJM: An International Journal of Medicine, 108(4), 273–277.

23 National Health Commission of the People’s Republic of China. Provisional rules of medical quality security incident report. (2011) August 2022, from http://www.nhc.gov.cn/wjw/gfxwj/201304/143166409ccf4232a71f97f203d25e05.shtml

24 Farley, D. O., Haviland, A., Champagne, S., Jain, A. K., Battles, J. B., Munier, W. B., & Loeb, J. M. (2008). Adverse-event-reporting practices by US hospitals: Results of a national survey. Quality and Safety in Health Care, 17(6), 416–423.

25 Yang J., Zhang J., Zhang Z., Zheng Z. (2016) Establishment of an internal reporting system of medical adverse events based on hospital informatization. Journal of Shanghai Jiaotong University(Medical Science), 2016,36(03): 423–426.

26 Ranaei, A., Gorji, H. A., Aryankhesal, A., & Langarizadeh, M. (2020). Investigation of medical error-reporting system and reporting status in Iran in 2019. Journal of Education and Health Promotion, 9.

27 Scott, S. D., Hirschinger, L. E., Cox, K. R., McCoig, M., Brandt, J., & Hall, L. W. (2009). The natural history of recovery for the healthcare provider “second victim” after adverse patient events. BMJ Quality & Safety, 18(5), 325–330.

28 Wu, A. W. (2000). Medical error: the second victim: the doctor who makes the mistake needs help too. Bmj, 320(7237), 726–727.

29 Yao SK, Zeng Q, Peng MQ, Ren SY, Chen G, Wang JJ. Stop violence against medical workers in China. J Thorac Dis. 2014;6(6):E141–E145.

30 Liu JL, Zheng J, Liu K, et al. Workplace violence against nurses, job satisfaction, burnout, and patient safety in Chinese hospitals. Nurs Outlook. 2019;67(5):558–566.

31 Savage, S. W., Schneider, P. J., & Pedersen, C. A. (2005). Utility of an online medication-error-reporting system. American journal of health-system pharmacy, 62(21), 2265–2270.

32 Cullen DJ, Bates DW, Small SD, Cooper JB, Nemeskal AR, Leape LL. The incident reporting system does not detect adverse drug events: A problem for quality improvement. Jt Comm J Qual Improv 1995;21:549–552.

33 Uribe CL, Schweikhart SB, Pathak DS, Dow M, Marsh GB. Perceived barriers to medical-error reporting: An exploratory investigation. J Healthc Manag 2002;47:263–279.

34 Kingston MJ, Evans SM, Smith BJ, Berry JG. Attitudes of doctors and nurses towards incident reporting: A qualitative analysis. Med J Aust 2004;181:36–39.

35 Schaufeli WB, Bakker AB, Hoogduin K, Schaap C, Kladler A. On the clinical validity of the maslach burnout inventory and the burnout measure. Psychol Health 2001;16:565–582.

36 Lee, H.-F., Yen, M., Fetzer, S., and Chien, T.W. (2015). Predictors of burnout among nurses in Taiwan. Community mental health journal 51, 733–737.

37 Ning, S., Zhong, H., Libo, W., & Qiujie, L. (2009). The impact of nurse empowerment on job satisfaction. Journal of advanced nursing, 65(12), 2642–2648.

38 Durham, M.E., Bush, P.W., and Ball, A.M. (2018). Evidence of burnout in health-system pharmacists. American Journal of Health-System Pharmacy 75, S93–S100.

39 Mészáros, V., Ádám, S., Szabó, M., Szigeti, R., and Urbán, R. (2014). The Bifactor Model of the Maslach Burnout Inventory–Human Services Survey (MBI□HSS)—An Alternative Measurement Model of Burnout. Stress and Health 30, 82–88.

40 Yang, J., Liu, Y., Chen, Y., & Pan, X. (2014). The effect of structural empowerment and organizational commitment on Chinese nurses’ job satisfaction. Applied Nursing Research, 27(3), 186–191.

41 Baron R, Kenny D. The Moderator-Mediator Variable Distinction in Social Psychological Research. Journal of personality and social psychology 1987;51:1173–82.

42 Sobel ME. Asymptotic Confidence Intervals for Indirect Effects in Structural Equation Models. Sociological Methodology 1982;13:290–312.

43 Carayon, P., Cassel, C., & Dzau, V. J. (2019). Improving the system to support clinician well-being and provide better patient care. JAMA, 322(22), 2165-2166.

44 Evans, S. M., Berry, J. G., Smith, B. J., Esterman, A., Selim, P., O’Shaughnessy, J., & DeWit, M. (2006). Attitudes and barriers to incident reporting: a collaborative hospital study. BMJ Quality & Safety, 15(1), 39–43.

45 Dyab, E. A., Elkalmi, R. M., Bux, S. H., & Jamshed, S. Q. (2018). Exploration of nurses’ knowledge, attitudes, and perceived barriers towards medication error reporting in a tertiary health care facility: A qualitative approach. Pharmacy, 6(4), 120.

46 Zhao, X., Zhao, S., Liu, N., & Liu, P. (2021). Willingness to report medical incidents in healthcare: a psychological model based on organizational trust and Benefit/Risk perceptions. The Journal of Behavioral Health Services & Research, 48(4), 583–596.

47 Chang IC, Hsu HM. Predicting medical staff intention to use an online reporting system with modified unified theory of acceptance and use of technology. Telemed J E Health. 2012 Jan-Feb;18(1):67–73.

48 Afolalu, O. O., Jordan, S., & Kyriacos, U. (2021). Medical error reporting among doctors and nurses in a Nigerian hospital: A cross□sectional survey. Journal of Nursing Management, 29(5), 1007–1015.

49 Lawton, R., & Parker, D. (2002). Barriers to incident reporting in a healthcare system. BMJ Quality & Safety, 11(1), 15–18.

50 Chiang, H. Y., Lee, H. F., Lin, S. Y., & Ma, S. C. (2019). Factors contributing to voluntariness of incident reporting among hospital nurses. Journal of nursing management, 27(4), 806–814.

51 Moumtzoglou, A. (2010). Factors impeding nurses from reporting adverse events. Journal of nursing management, 18(5), 542–547.

52 Vrbnjak, D., Denieffe, S., O’Gorman, C., & Pajnkihar, M. (2016). Barriers to reporting medication errors and near misses among nurses: A systematic review. International journal of nursing studies, 63, 162–178.

53 Ramírez, E., Martín, A., Villán, Y., Lorente, M., Ojeda, J., Moro, M., … & Frank, A. (2018). Effectiveness and limitations of an incident-reporting system analyzed by local clinical safety leaders in a tertiary hospital: Prospective evaluation through real-time observations of patient safety incidents. Medicine, 97(38).

54 Savage, S. W., Schneider, P. J., & Pedersen, C. A. (2005). Utility of an online medication-error-reporting system. American journal of health-system pharmacy, 62(21), 2265–2270.

55 Costello, J. L., Torowicz, D. L., & Yeh, T. S. (2007). Effects of a pharmacist-led pediatrics medication safety team on medication-error reporting. American journal of health-system pharmacy, 64(13), 1422–1426.

56 Carthon, J. M. B., Hatfield, L., Brom, H., Houton, M., Kelly-Hellyer, E., Schlak, A., & Aiken, L. (2021). System-level improvements in work environments lead to lower nurse burnout and higher patient satisfaction. Journal of nursing care quality, 36(1), 7.

57 Wu, J.-H. et al., 2007. Testing the technology acceptance model for evaluating healthcare professionals’ intention to use an adverse event reporting system. International Journal for Quality in Health Care, 20(2), pp.123–129.

58 Cullen, D. J., Bates, D. W., Small, S. D., Cooper, J. B., Nemeskal, A. R., & Leape, L. L. (1995). The incident reporting system does not detect adverse drug events: a problem for quality improvement. The Joint Commission journal on quality improvement, 21(10), 541–548.

59 Lewis, E. J., Baernholdt, M., & Hamric, A. B. (2013). Nurses’ experience of medical errors: An integrative literature review. Journal of Nursing Care Quality, 28(2), 153–161.

60 Lewis, E. J., Baernholdt, M., & Hamric, A. B. (2013). Nurses’ experience of medical errors: An integrative literature review. Journal of Nursing Care Quality, 28(2), 153–161.

61 Kalisch, B., Tschannen, D., & Lee, H. (2011). Does missed nursing care predict job satisfaction?. Journal of Healthcare Management, 56(2), 117–134.

62 Davis K, Doty MM, Shea K, Stremikis K. Health information technology and physician perceptions of quality of care and satisfaction. Health Policy. 2009;90(2-3):239–246.

63 Vogus, T. J., Ramanujam, R., Novikov, Z., Venkataramani, V., & Tangirala, S. (2020). Adverse events and burnout: the moderating effects of workgroup identification and safety climate. Medical care, 58(7), 594–600.

64 Weingart, S. N., Pagovich, O., Sands, D. Z., Li, J. M., Aronson, M. D., Davis, R. B., … & Phillips, R. S. (2005). What can hospitalized patients tell us about adverse events? Learning from patient□reported incidents. Journal of general internal medicine, 20(9), 830–836.

65 Gowen, C.R., McFadden, K.L. and Settaluri, S. (2012), Contrasting continuous quality improvement, Six Sigma, and lean management for enhanced outcomes in US hospitals, American Journal of Business, Vol. 27 No. 2, pp. 133–153.

66 Zeng, J.-Y., An, F.-R., Xiang, Y.-T., Qi, Y.-K., Ungvari, G. S., Newhouse, R., et al. (2013). Frequency and risk factors of workplace violence on psychiatric nurses and its impact on their quality of life in China. Psychiatry Research 210, 510–514. doi:10.1016/j.psychres.2013.06.013.

67 Stavropoulou, C., Doherty, C., & Tosey, P. (2015). How effective are incident□reporting systems for improving patient safety? A systematic literature review. The Milbank Quarterly, 93(4), 826–866.

68 Tan, C. N. L., Ojo, A. O., Cheah, J. H., & Ramayah, T. (2019). Measuring the influence of service quality on patient satisfaction in Malaysia. Quality Management Journal, 26(3), 129–143.

69 Galián-Muñoz, I., Ruiz-Hernández, J. A., Llor-Esteban, B., and López-García, C. (2014). User violence and nursing staff Burnout. Journal of Interpersonal Violence 31, 302–315. doi:10.1177/0886260514555367.

70 Zimmer KK, Cabelus NB (2003) Psychological effects of violence on forensic nurses. J Psychosoc Nurs Ment Health Serv 41(11): 28–35

71 Liu, J., Zheng, J., Liu, K., Liu, X., Wu, Y., Wang, J., & You, L. (2019). Workplace violence against nurses, job satisfaction, burnout, and patient safety in Chinese hospitals. Nursing outlook, 67(5), 558–566.

72 Gleeson L, Dalton K, O’Mahony D, Byrne S. Interventions to improve reporting of medication errors in hospitals: A systematic review and narrative synthesis. Res Social Adm Pharm. 2020 Aug;16(8):1017–1025.

73 Force, M. V., Deering, L., Hubbe, J., Andersen, M., Hagemann, B., Cooper-Hahn, M., & Peters, W. (2006). Effective strategies to increase reporting of medication errors in hospitals. Jona: the Journal of Nursing Administration, 36(1), 34–41.

74 Nakajima, K., Kurata, Y., & Takeda, H. (2005). A web-based incident reporting system and multidisciplinary collaborative projects for patient safety in a Japanese hospital. BMJ Quality & Safety, 14(2), 123–129.

